# UniTox: Leveraging LLMs to Curate a Unified Dataset of Drug-Induced Toxicity from FDA Labels

**DOI:** 10.1101/2024.06.21.24309315

**Authors:** Jake Silberg, Kyle Swanson, Elana Simon, Angela Zhang, Zaniar Ghazizadeh, Scott Ogden, Hisham Hamadeh, James Zou

## Abstract

Drug-induced toxicity is one of the leading reasons new drugs fail clinical trials. Machine learning models that predict drug toxicity from molecular structure could help researchers prioritize less toxic drug candidates. However, current toxicity datasets are typically small and limited to a single organ system (e.g., cardio, renal, or liver). Creating these datasets often involved time-intensive expert curation by parsing drug label documents that can exceed 100 pages per drug. Here, we introduce UniTox^1^, a unified dataset of 2,418 FDA-approved drugs with drug-induced toxicity summaries and ratings created by using GPT-4o to process FDA drug labels. UniTox spans eight types of toxicity: cardiotoxicity, liver toxicity, renal toxicity, pulmonary toxicity, hematological toxicity, dermatological toxicity, ototoxicity, and infertility. This is, to the best of our knowledge, the largest such systematic human *in vivo* database by number of drugs and toxicities, and the first covering nearly all FDA-approved medications for several of these toxicities. We recruited clinicians to validate a random sample of our GPT-4o annotated toxicities, and UniTox’s toxicity ratings concord with clinician labelers 87–96% of the time. Finally, we benchmark a graph neural network trained on UniTox to demonstrate the utility of this dataset for building molecular toxicity prediction models.

## 1 Introduction

An estimated 90% of drugs fail in clinical trials [1]. While the most common cause of failure is efficacy, one study found that the second largest cause (24% of failures) was drug safety [2]. Further, every year, previously approved drugs are taken off the market as unanticipated toxicities become apparent in post-marketing data that can be difficult to screen pre-clinically [3]. These different drug-induced toxicities span many different organ systems, including the heart, liver, kidneys, blood, and lungs. As a result, there is a strong need for predictive models that can anticipate a broad range of human *in vivo* toxicities so that researchers can screen for molecules with the highest chance of clinical trial and post-market safety and success.

A major source of both data and expertise in evaluating drug-induced toxicity is the FDA. FDA researchers have published analyses of FDA-approved drug labels on drug-induced cardiotoxicity (DICTrank [4]), drug-induced liver injury (DILIrank [5]), and drug-induced renal toxicity (DIRIL [6]). Each analysis has involved one or more trained professionals who carefully comb through each label to make a toxicity determination. More recently, the FDA has explored the use of large language models (LLMs) to process drug labels more quickly [7]. They developed askFDALabel, a retrieval-augmented generation (RAG) [8] system that finds the most similar label fragments to a user query, then utilizes a fine-tuned LLM to generate a response based on those fragments. They showcase askFDALabel for assessing drug-induced cardiotoxicity (DICT) and find that, where labels were available, askFDALabel agrees with the human-labeled dataset 78% of the time.

In addition to that work, several other toxicity databases have been developed. For example, Cavasotto and Scardino [9] compiled a set of toxicity databases. These existing datasets have several limitations. First, these datasets are often small due to time-consuming labeling efforts [6]. Second, these datasets use different methodologies to evaluate toxicities. For example, the FDA’s DIRIL (renal toxicity) work draws on two existing datasets that disagreed more than 30% of the time on the same drugs [10, 11]. Many of these, such as SIDER [12], ECHA’s C&L system [13], PubChem’s Hazardous Substances Data Bank [14], and the Comparative Toxicogenomics Database [15], cannot be used to search by toxicity status and do not include all toxicity keywords in their side effects or phenotype data. While Tox21 and ToxCast [16] cover a large number of chemicals, not limited to FDA-approved medications, they are based on *in vitro* assays that may not accurately reflect *in vivo* drug effects. These chemical databases also typically exclude biologics. Other very comprehensive toxicity databases, such as PNEUMOTOX [17] for pulmonary toxicity and LiverTox [18] for liver toxicity, cover only a single organ system and may differ in methodologies. Machine learning models for toxicity that are trained on these datasets [19, 20, 21, 22, 23], while useful, suffer from the same limitations as the underlying datasets.

### Our contributions

In this work, we develop a framework for using LLMs to rapidly categorize the toxicity of drugs from FDA labels. We apply this methodology to build UniTox, the largest human *in vivo* cross-toxicity dataset of 2,418 FDA-approved drugs. We evaluate the accuracy of these predictions, achieving up to 93% accuracy on pre-existing datasets compared to 78% for askFDALabel, and as well as up to 87–96% concordance on a clinician-reviewed sample. Finally, we benchmark the performance of a graph neural network trained on small molecule drugs from UniTox to illustrate the benefit of building a uniform toxicity dataset.

## 2 Methods

### 2.1 Building UniTox

To build UniTox, we first needed to curate a set of drugs and associated drug labels to analyze. Drawing inspiration from askFDALabel, we started with the universe of all human prescription drugs from the FDALabel database [24]. One important difference from askFDALabel is that we included biologic drugs, as those were included in DICTrank. We then grouped drugs by unique generic drug names and removed labels where the route of administration included topical, irrigational, or intradermal. For each unique generic drug name where we had an exact label match with askFDALabel, we used the same label. Where we did not have an exact match, we used the most recent New Drug Application (NDA) label for that generic drug name. Where we did not have an NDA label, we used the most recent Abbreviated New Drug Application (ANDA) label, which is used for generic versions of brand-name drugs.

This process, outlined in Figure 1, gave us a set of 2,418 drugs and drug labels for UniTox. Then, we applied our LLM framework to the UniTox drugs for each of eight types of toxicity: cardiotoxicity, liver toxicity, renal toxicity, pulmonary toxicity, hematological toxicity, dermatological toxicity, ototoxicity, and infertility.

**Figure 1:**
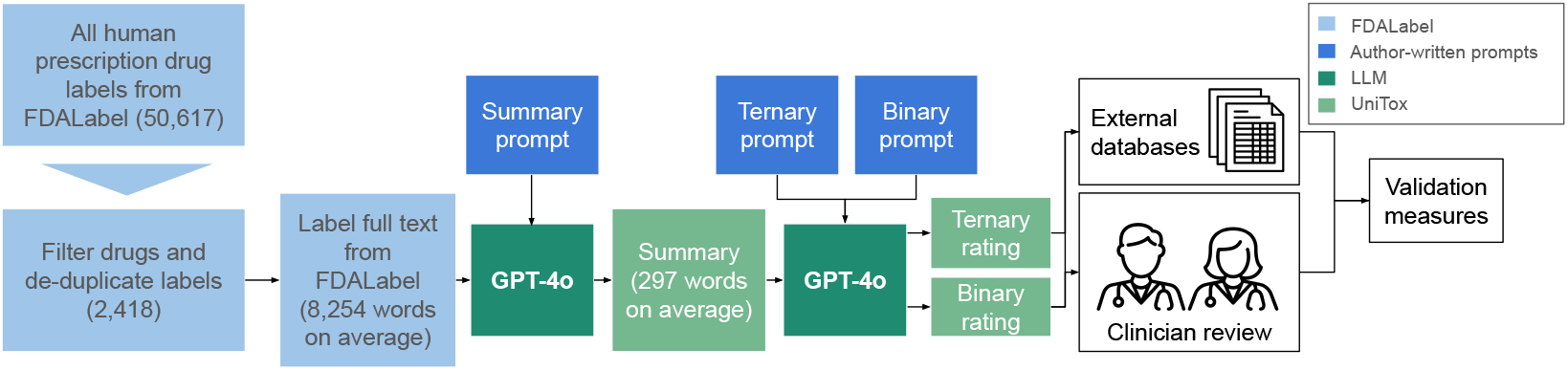
UniTox was built by applying a large language model (GPT-4o) to a curated set of 2,418 FDA drug labels to produce ternary (No/Less/Most) and binary (No/Yes) toxicity ratings, which were evaluated based on external databases and clinican review.

### 2.2 Generating toxicity ratings with LLMs

To generate toxicity ratings from a drug label, we utilized an LLM and chain-of-thought [25] reasoning with a two-tiered prompt system. The first prompt—the “summary prompt”—asks the LLM to read the drug label and summarize the drug’s toxicity for a given type of toxicity (e.g., cardiotoxicity). The second prompt—the “rating prompt”—asks the LLM to use only this toxicity summary to produce a toxicity rating, which is either a ternary rating (No, Less, or Most toxicity) or a binary rating (No or Yes toxicity). We anticipated that the “Less” category of the ternary rating system can function as a proxy of the model’s confidence, with “No” and “Most” as more confident predictions, so we focus on the ternary rating system in our results. The specific prompts provided to the model are below, where `<toxicity type>` is replaced with the toxicity type.

**Summary Prompt**

Provide a summary of all the parts of the drug label that discuss `<toxicity type>` risks and `<toxicity type>` reactions for this drug. In your summary of each sentence, clearly state whether the drug itself was associated with or caused the `<toxicity type>` risk.

**RatRating Prompt – Ternary**

Given the above information about a drug, answer “was this drug associated with No `<toxicity type>`, Less `<toxicity type>`, or Most `<toxicity type>`?” Now, answer with just one word: No, Less or Most.

**Rating Prompt – Binary**

Given the above information about a drug, answer “was this drug associated with `<toxicity type>`?” Now, answer with just one word: Yes or No.

### 2.3 Validation on DICTrank, DILIrank, and DIRIL

We first validated the toxicity ratings in UniTox by measuring the concordance of these ratings with human-labeled toxicity ratings from three FDA-designed datasets: DICTrank, DILIrank, and DIRIL. This required matching the drugs in UniTox to those in the FDA datasets using the drug data available in these datasets. For DICTrank, we matched by generic drug name. For DILIrank, we used the RxNorm [26] database to pull Structured Product Labeling (SPL) Set IDs for each drug, then matched to the SPL IDs we used. For DIRIL, we matched to our toxicity generations on moiety UNII codes. Then, for each of these three datasets, we evaluated UniTox and human toxicity rating concordance among the matched drugs. Furthermore, to better understand what drives the LLM’s performance, we performed ablations on DICTrank in Section 3.2.1, including a longer prompt with the specific cardiotoxic keywords from DICTrank, using GPT-3.5 instead of GPT-4o, and removing the chain-of-thought step.

### 2.4 Clinician validation on other toxicities

For the five remaining toxicity types without pre-existing validation data, we worked with clinicians to manually validate a subset of the UniTox toxicity ratings. Specifically, we asked clinicians to read the toxicity summary and use both the summary and their knowledge of the drug to validate the toxicity ratings for 100 randomly sampled drugs for each of the five toxicity types (two clinicians, 50 drugs per clinician per toxicity type). For each drug and toxicity type, the clinicians separately evaluated both the ternary and binary toxicity ratings on a scale of 1 to 3, where 1 means “The model’s score is factually correct and I agree with it”, 2 means”The model’s score is reasonable but I don’t necessarily agree with it completely”, and 3 means “The model’s score is factually incorrect and I disagree with it”. We also asked clinicians to flag if the LLM-generated toxicity summary did not concord with their understanding of a drug and its use.

## 3 Results

Here, we present details about the UniTox dataset (Section 3.1). Then, we discuss our validations on external datasets and the effect of ablations on performance (Section 3.2). Next, we show results of our clinician review of the five toxicities without pre-existing validation data (Section 3.3). Finally, we illustrate the benefit of a unified toxicity dataset by training a GNN on UniTox (Section 3.4).

### 3.1 UniTox

UniTox contains 2,418 drugs with eight types of toxicities. For each drug and toxicity type, UniTox includes (1) a GPT-4o generated summary of the drug label’s discussion of that toxicity, (2) a ternary classification into No Toxicity, Less Toxicity, or Most Toxicity, (3) a binary classification into No Toxicity and Yes Toxicity, and (4) the Stuctured Product Labeling (SPL) ID for the document used to generate all data. Properties 1-3 are listed for two examples in Table 1. A key value of UniTox lies in its summaries, which capture the nuance of each drug’s toxicity in a fraction of the length of the full text labels (297 words on average in the summary compared to 8,254 words on average in the full label). The value also lies in the toxicity ratings, which can be used as labels for training downstream toxicity predictors. Where users wish to modify our LLM-generated ratings, they can utilize the short summaries and avoid the need to read full-text drug labels.

**Table 1:** Example UniTox Entries.

|  |  |  |
| --- | --- | --- |
| Generic Name | ABALOPARATIDE | ABEMACICLIB |
| Toxicity | Pulmonary | Pulmonary |
| Ternary Rating | No | Most |
| Binary Rating | No | Yes |
| Summary (Trimmed) | ...The sections of the label that detail adverse reactions, warnings, and precautions do not mention any pulmonary-related issues directly associated... | ... associated with significant pulmonary toxicity risks, including severe, life-threatening, or fatal interstitial lung disease (ILD) or pneumonitis... observed in clinical trials, postmarketing settings... |

UniTox is, to the best of our knowledge, the largest human *in vivo* drug-induced toxicity database by number of drugs and number of toxicities. It covers a diverse range of drugs and clinical toxicities that can often be difficult to identify in pre-clinical studies, containing both positive and negative examples of drug toxicities. Figure 2 shows the number of drugs in UniTox with each ternary toxicity rating for each toxicity type. While most toxicity types have a balance of toxic and non-toxic drugs (30–65% classified as Most Toxic), it is worth noting that dermatological toxicity and ototoxicity are outliers with 85% and 8% of the drugs predicted as Most Toxic, respectively.

**Figure 2:**
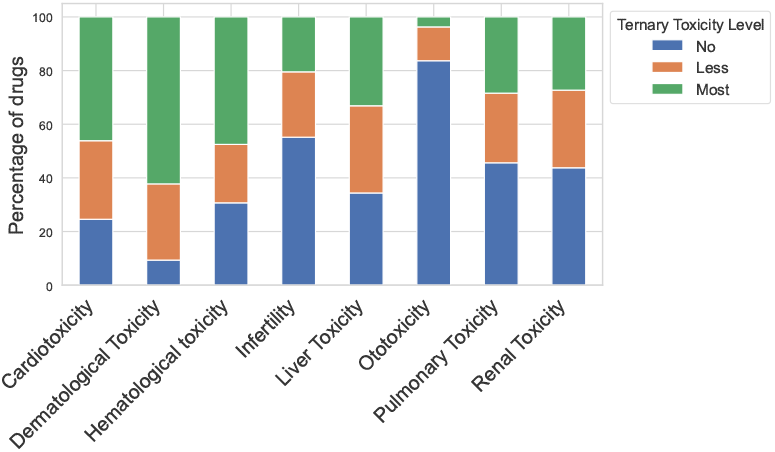
Distribution of ternary toxicity ratings in UniTox across 2,418 drugs.

#### 3.1.1 Cross-toxicity analysis

One of the advantages of a unified toxicity dataset is the ability to determine whether drugs exhibit multiple toxicities. This is, to the best of our knowledge, the largest systematic analysis across drug classes of how different drug toxicities are related. Interestingly, we find that the number of toxicities per drug approximates a normal distribution, centered at four of the eight toxicities.

We then calculated pairwise correlations across the toxicities, using our binary ratings (Figure 3). We find that liver toxicity and hematological toxicity are the most highly correlated, at 0.45, with pulmonary and cardiotoxicity the second most correlated at 0.30, and liver toxicity and renal toxicity third most correlated at 0.29. We did not find any negative correlations. We believe these results can help future researchers better understand drug toxicity by examining potential causes of these correlations and specific drugs that exhibit unusual patterns of toxicity across systems.

**Figure 3:**
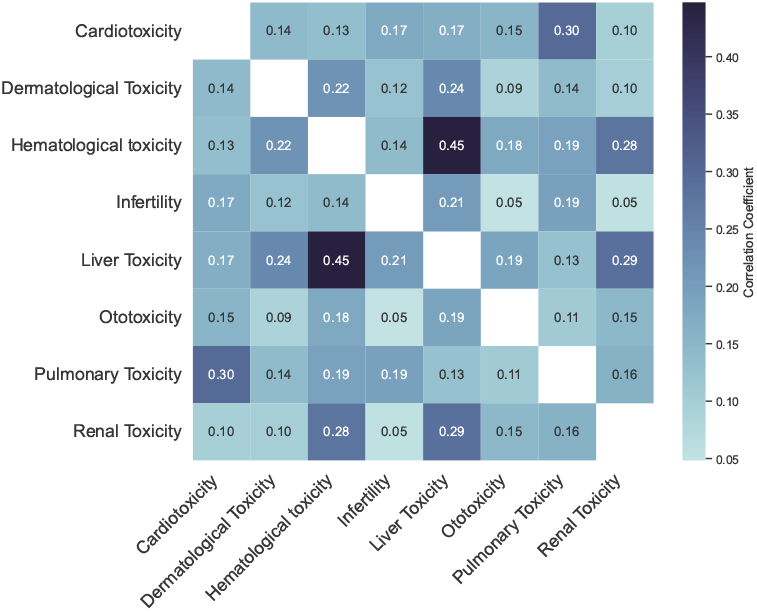
Heatmap of correlations between different toxicity types in UniTox.

### 3.2 Validation on external datasets

#### 3.2.1 DICTrank

UniTox has 1,181 label matches with the DICTrank dataset of 1,318 drugs. Usually, a lack of a match indicates the drug has been withdrawn or discontinued and so a label is no longer available. To binarize our results, we consider “Ambiguous-DICT-Concern”, “Less-DICT-Concern”, and “Most-DICT-Concern” to be toxic, and “No-DICT-Concern” to be non-toxic. We similarly binarized our ternary ratings by combining “Less” and “Most” into a single toxic category. This is the **Ternary** column in Table 2. We also show results from dropping the “Less” category to focus on high confidence predictions, in the **Ternary w/o Less** column. Finally, to match askFDALabel, we show the results of binary ratings on only the ground truth “No-DICT-Concern” and “Most-DICT-Concern” subset (**Binary on GT No/Most** column). Here, we obtain a significantly improved 93.0% accuracy compared to askFDALabel’s 77.7% accuracy with a fine-tuned LLM and 71.5% with GPT-3.5. We also present negative and positive predictive values as we want to maximize the share of each predicted class that, if used as training labels, would have the correct labels.

**Table 2:** DICTrank, DILIrank, and DIRIL validation results.

| DICTrank | Ternary | Ternary w/o Less (n=761) | Binary on GT No/Most |
| --- | --- | --- | --- |
| Accuracy (%) | 84.6 | 92.5 | 93.0 |
| NPV (%) | 79.6 | 79.7 | 96.2 |
| PPV (%) | 85.7 | 97.1 | 90.0 |
| DICT ablation accuracies | Ternary | Ternary w/o Less (n) | Binary on GT No/Most |
| Keyword summary prompt (%) | 88.1 | 90.4 (924) | 93.7 |
| No CoT (%) | 77.8 | 67.4 (629) | 92.5 |
| GPT-3.5 (%) | 77.6 | 77.8 (855) | 88.4 |
| RAG fragment context (%) |  |  | 94.2 |
| askFDALabel (previous SOTA) |  |  | 77.7 |
| DILrank | Ternary | Ternary w/o Less (n=525) | Binary on GT No/Most |
| Accuracy (%) | 81.1 | 85.0 | 86.2 |
| NPV (%) | 72.4 | 72.4 | 97.5 |
| PPV (%) | 83.8 | 92.4 | 71.9 |
| DIRIL | Ternary | Ternary w/o Less (n=171) | Binary on GT No/Most |
| Accuracy (%) | 71.3 | 76.8 | 72.9 |
| NPV (%) | 75.9 | 75.9 | 66.4 |
| PPV (%) | 69.4 | 77.7 | 80.2 |

#### 3.2.2 DICTrank ablations and sensitivity analysis

To better understand our performance on the DICTrank dataset, we consider a series of ablations (Table 2). First, we used a keyword summary prompt that contained the full list of DICTrank keywords (e.g., myocardial infarction and Torsade de Pointes). We did not alter the ratings prompts. Performance increases or decreases slightly on our different cuts of the data, likely demonstrating that the GPT-4o model has a strong and accurate internal definition of cardiotoxicity.

Second, we ablated the chain-of-thought step (i.e., the summary prompt), instead providing the full text of the drug label to the model when using the ternary and binary ratings prompts. We note a consistent decrease in performance. Considering that our base ratings had access to only the summaries, this shows the benefit of providing focused and thoughtful information about toxicity.

Third, we switched to GPT-3.5, which required truncating a small number of labels to fit into context. askFDALabel achieved a DICTrank accuracy of 71.5% using GPT-3.5 while our use of GPT-3.5, which applies our prompting strategy, achieved 88.4% accuracy. In particular, our prompt specifically asked about “cardiotoxicity” while askFDALabel asked about “cardio-related adverse events or risks”, which likely boosted performance. GPT-3.5 consistently performs worse than GPT-4o.

Finally, to better understand the role of using the full drug label, we applied our ratings prompts on the RAG-retrieved label fragments from askFDALabel. Both performed similarly. However, generating GPT-4o summaries only involved designing a short prompt, compared to building a custom RAG system, and should cover all relevant information in the drug label. The RAG system returns only the top-k fragments, so it may miss vital details. For example, our summary of the full label of voclosporin (below) discussed the risk of QT prolongation in several sections of the label that were not returned by the RAG system. As a result, only the prediction based on full context was correct.

**Voclosporin: GPT-4o cardiotoxicity summary** | **DICTrank: Toxic** | **GPT-4o: Toxic**

…

**4. Medication Guide, What are the possible side effects of LUPKYNIS? (Section 17): Summary:** LUPKYNIS may cause serious side effects, including a serious heart rhythm problem (QT prolongation).

**Association**: The drug itself is associated with the risk of a serious heart rhythm problem (QT prolongation).

In summary, LUPKYNIS is associated with cardiotoxicity risks, specifically QT prolongation, which is dose-dependent and can be clinically significant when combined with other QT-prolonging drugs. This risk is highlighted in multiple sections of the drug label, indicating that the drug itself can cause these cardiotoxic effects.

Most interestingly, our GPT-4o-generated summaries can reason through difficult edge cases. For example, linagliptin is rated as Most-DICT-Concern for cardiotoxicity in DICTrank as the label discusses heart failure. However, the label notes that “Heart failure has been observed with two other members of the DPP-4 inhibitor class.” GPT-4o, looking only at the three results returned by the RAG system, predicts Yes for cardiotoxicity. However, GPT-4o provided with the full drug label notes lingagliptin itself has not been associated with heart failure and discusses two large-scale trials showing linagliptin did not increase major adverse cardiovascular events (see box below). As a result, it predicts linagliptin has No Toxicity in the binary case. While this differs from the ground truth in DICTrank, it is a reasonable response that comprehensively reasons across the drug label.

**Linagliptin:** GPT-4o cardiotoxicity summary | DICTrank: Toxic | GPT-4o: Non-Toxic

…

**Observation**: Heart failure has been observed with two other members of the DPP-4 inhibitor class.

**Association** It is not explicitly stated that TRADJENTA itself causes heart failure, but the risk is considered due to the class effect.

…

**CARMELINA Trial**: **Primary Endpoint**: The primary endpoint was the time to first occurrence of major adverse cardiovascular events (MACE), including cardiovascular death, non-fatal myocardial infarction, or non-fatal stroke. **Results**: The estimated hazard ratio for MACE associated with TRADJENTA relative to placebo was 1.02, indicating no significant increase in risk. **Conclusion**: TRADJENTA did not show an increased risk of major adverse cardiovascular events compared to placebo.

…

**Conclusion: Heart Failure**: While heart failure has been observed with other DPP-4 inhibitors, TRADJENTA itself is not explicitly stated to cause heart failure but should be used with caution in patients with risk factors. **Cardiovascular Events**: Clinical trials (CARMELINA and CAROLINA) indicate that TRADJENTA does not increase the risk of major adverse cardiovascular events compared to placebo or glimepiride.

…

#### 3.2.3 DILIrank and DIRIL

We performed similar validations of DILIrank (liver toxicity) and DIRIL (renal toxicity), as seen in Table 2. For DILIrank, we achieve similar performance as DICTrank on the 819 drugs where we had a match. Our DIRIL results are less impressive, which may be due to the fact that DIRIL was constructed using a different methodology compared to DICTrank and DILIrank, in which they took labels from previous papers as given instead of primarily using FDA labels. For example, we note that for 9 of our 25 false positives and for 8 of our 48 false negatives, at least one of the previous papers agreed with GPT-4o’s rating rather than the FDA paper’s determination.

However, it is also likely that GPT-4o’s internal definition of renal toxicity is less calibrated to the FDA’s definition than for other toxicities. Given that false negatives were more frequent in this case than false positives, we analyzed the false negative cases and found that the GPT-4o-generated summary often noted that the use of the drug is cautioned in renally impaired patients. When binarizing this summary, GPT-4o generated No Toxicity in these cases when the FDA reviewers likely viewed this as a sign of toxicity. This shows that, even when the binary rating of the model may differ from human labels, GPT-4o condenses valuable information for human reviewers.

### 3.3 Clinician evaluation of toxicity ratings

Figure 4 shows the distribution of clinician-derived scores of the LLM-generated UniTox labels (ternary rating). Depending on the toxicity, 87-96% of the drugs were considered accurately labeled, 3-12% were ambiguous, and 1-12% were labeled incorrect.

**Figure 4:**
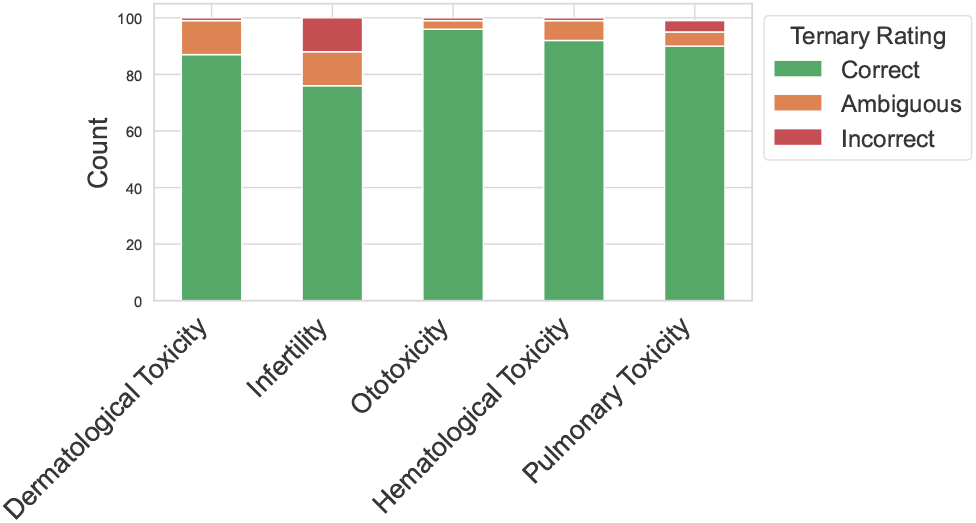
Distribution of clinician validation scores on GPT-4o-generated ternary toxicity ratings.

While at least 87% of the clinician scores agreed with the UniTox labels, the disagreements reveal some edge cases. For many of the drugs where the clinicians gave a rating of 2, the explanation was a lack of direct data or evidence in humans for the specific toxicity. For example, trilaciclib got a UniTox rating of Most Toxicity due to evidence that it may impair fertility in animals; however, the clinician scored this rating with a 2 due to the lack of *human* evidence. On the other hand, trientine hydrochloride capsules got a UniTox rating of No Toxicity as the label provided zero evidence that this drug is associated with fertility risks; this also received a clinician score of 2 as the label simply did not provide any data about fertility risks and the model was conflating a lack of evidence about toxicity with evidence for a lack of toxicity. Indeed, based on this feedback, we added a new prompt for infertility to clarify the level of available evidence (added as an additional column in UniTox).

A few of the validation set examples where clinicians disagreed with the model highlight genuine errors in the model’s assessment. For example, ganciclovir injection got a UniTox rating of Less Toxicity for ototoxicity. However, the label explicitly lists “tinnitus, ear pain, deafness” as observed adverse effects, which should clearly be considered Most Toxicity.

### 3.4 GNN toxicity prediction

Next, we demonstrate the utility of UniTox by using it to train machine learning models to predict toxicity from molecular structure, which is an important aspect of drug discovery. We trained a widely-used Chemprop-RDKit model, which is a Chemprop [19] graph neural network augmented with 200 molecular features computed by the cheminformatics package RDKit [27]. We performed ten-fold cross-validation using a challenging scaffold split, which means that molecules were clustered by their core molecular scaffold and clusters were placed either entirely in the train set or entirely in the test set. This ensures that similar drugs do not leak between train and test. The Chemprop-RDKit model is trained in a multi-task setting with one model predicting all eight toxicities.

Since Chemprop-RDKit is only designed to work with small molecules, we restricted UniTox to the set of small molecule drugs (e.g., excluding biologics). We then used the PubChem [28] API to match generic drug names to SMILES. We deduplicated drugs by SMILES and removed any SMILES where at least one of the toxicity ratings across the eight toxicities differ between different drugs with the same SMILES (e.g., different formulations of the same drug). This resulted in a deduplicated set of 1,349 drugs with unique SMILES and concordant toxicity ratings, which we refer to as the UniTox-GNN subset. We trained our models in two binary classification settings: (1) binary, where the GNN simply predicts the binary rating, and (2) confident ternary, where the GNN predicts No Toxicity or Most Toxicity and ignores Less Toxicity from the ternary ratings.

As shown in Figure 5, the Chemprop-RDKit model performs reasonably well, with mean ROC-AUCs exceeding 0.7 on five of the eight datasets in the confident ternary setting. Given the dataset size and the inherent biological complexity of human *in vivo* toxicity, this performance is reasonable and is within the range of other molecular property prediction models in the literature (e.g., ADMET-AI [20]). While the model has poor performance on dermatological toxicity and ototoxicity, this is likely due to the extreme class imbalance present in both datasets (87% with dermatological toxicity and 5% with ototoxicity in the UniTox-GNN ternary ratings). Overall, these results illustrate the benefit of building comprehensive toxicity datasets as it enables training molecular property prediction models that can generalize to new molecules and could potentially be used as an *in silico* toxicity screening tool prior to clinical validation.

**Figure 5:**
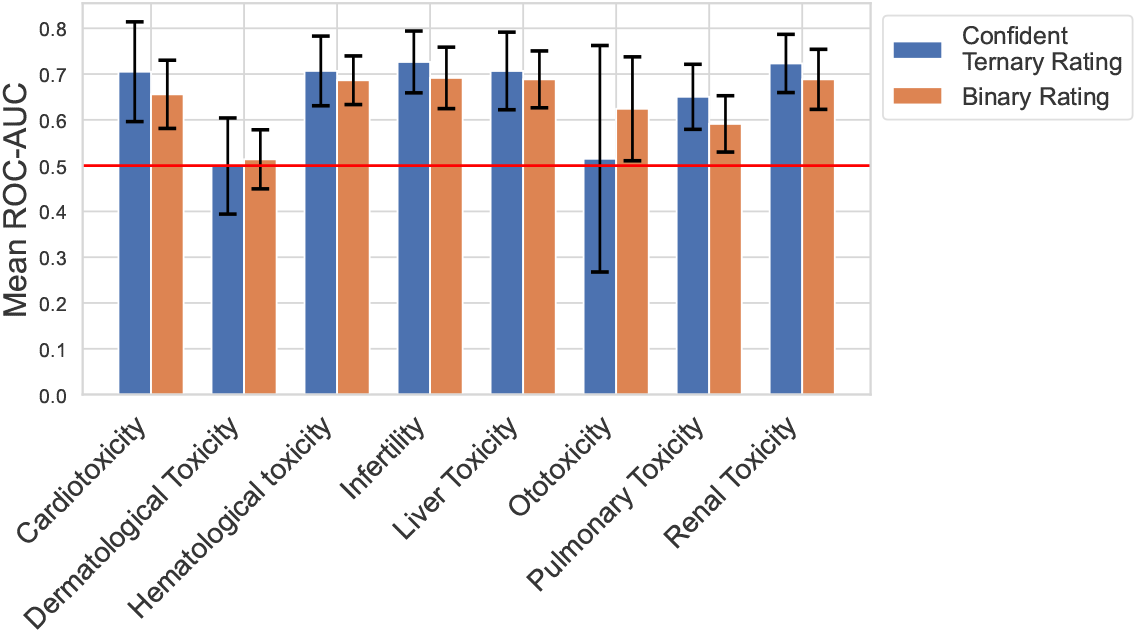
Performance of Chemprop-RDKit trained on the UniTox-GNN subset using either confident ternary or binary ratings (mean *±* standard deviation ROC-AUC across ten-fold cross-validation).

## 4 Discussion

In this work, we demonstrated the ability of GPT-4o to rapidly generate useful and accurate summaries of complex drug labels. When binarized, these summaries had high concordance with the external DICTrank (cardiotoxicity) and DILIrank (liver toxicity) datasets, and to a lesser extent, to the DIRIL (renal toxicity) dataset. UniTox also had a high concordance with clinical reviewers, even for toxicities without pre-existing comparable quantitative validation data. We demonstrate the value of these summaries, and their binarized values, by training molecular classifiers with predictive value in most cases. These labels, even where occasionally noisy, can serve as a benchmark for future classifiers that seek to demonstrate consistent performance across toxicities. Such consistent evaluation of downstream classifiers was not previously possible. Finally, we provide insight into the co-occurrence of multiple toxicities from drugs in a unified format not previously available.

The clearest limitation of our work is the challenge of going from a nuanced summary of the drug label to a binary or ternary rating. We note that common challenges are how to binarize toxicity in cases where (1) toxicity occurred only in specific or pre-disposed populations (e.g., children or impaired patients), (2) toxicity occurred only in other drugs of the same class, only in animals, or only at high doses that may exceed clinical relevance, (3) common but mild reactions (e.g., rashes for dermatological toxicity), and (4) rare reactions not easily observed except when specifically studied (e.g., infertility), in which case a lack of evidence may not be sufficient to conclude a lack of toxicity. These circumstances were often discussed in detail in GPT-4o’s generated summaries but were lost in the binary or ternary ratings. While we preferred consistent and simple prompts, perhaps more complicated, guided ratings prompts could better handle these. For example, future work could set a higher bar for dermatological toxicity.

Additionally, we note the ethical importance of accuracy in this case. We have taken steps to validate our predictions, and we provide the nuanced GPT-4o summaries based on drug labels. Still, we note here that these are LLM-generated predictions intended for drug research; they are not medical advice and are not meant to inform healthcare decisions.

As LLMs become more frequently used in information extraction tasks, it is important to understand their strengths and limitations. We demonstrate their value by creating an accurate and useful dataset in a fraction of the time it would take humans to process this massive amount of text. In particular, we show their ability to summarize text while maintaining its key information and nuance, and we create useful labels for downstream classifiers. We also highlight particular reasons why further condensing that information content into a single word remains challenging, for both models and humans.

## Data Availability

All data produced are available online at https://zou-group.github.io/UniTox-website/

## Footnotes

1 UniTox data is available at https://zou-group.github.io/UniTox-website. Code available at: https://github.com/jsilbergDS/UniTox

